# In-hospital and 30-day mortality after percutaneous coronary intervention in England before and after the COVID-19 era

**DOI:** 10.1101/2020.07.18.20155549

**Authors:** Mohamed O. Mohamed, Tim Kinnaird, Nick Curzen, Peter Ludman, Jianhua Wu, Muhammad Rashid, Ahmad Shoaib, Mark de Belder, John Deanfield, Chris P Gale, Mamas A. Mamas

**Affiliations:** Keele Cardiovascular Research Group, Centre for Prognosis Research, Keele University, United Kingdom; Department of Cardiology, Royal Stoke University Hospital, Stoke-on-Trent, United Kingdom; Department of cardiology, University hospital of Wales, Cardiff, Wales, UK; Wessex Cardiothoracic Unit, Southampton University Hospital Southampton & Faculty of Medicine University of Southampton, UK; Institute of Cardiovascular Sciences, University of Birmingham, Birmingham, UK; Leeds Institute for Data analytics, University of Leeds, Leeds, UK; National Institute for Cardiovascular Outcomes Research, Barts Health NHS Trust, London, UK; Institute of Cardiovascular Sciences, University College London, UK; Leeds Institute of Cardiovascular and Metabolic Medicine, University of Leeds, Leeds, UK; Department of Cardiology, Leeds Teaching Hospitals NHS Trust, Leeds, UK

**Keywords:** COVID-19, percutaneous coronary intervention, deaths, England, outcomes

## Abstract

**Objectives:** To examine short-term primary causes of death after percutaneous coronary intervention (PCI) in a national cohort before and during COVID-19.

**Background:** Public reporting of PCI outcomes is a performance metric and a requirement in many healthcare systems. There are inconsistent data on the causes of death after PCI, and what proportion of these are attributable to cardiac causes.

**Methods:** All patients undergoing PCI in England between 1^st^ January 2017 and 10^th^ May 2020 were retrospectively analysed (n=273,141), according to their outcome from the date of PCI; no death and in-hospital, post-discharge, and 30-day death.

**Results:** The overall rates of in-hospital and 30-day death were 1.9% and 2.8%, respectively. The rate of 30-day death declined between 2017 (2.9%) and February 2020 (2.5%), mainly due to lower in-hospital death (2.1% vs. 1.5%), before rising again from 1st March 2020 (3.2%) due to higher rates of post-discharge mortality. Only 59.6% of 30-day deaths were due to cardiac causes, the most common being acute coronary syndrome, cardiogenic shock and heart failure, and this persisted throughout the study period. 10.4% of 30-day deaths after 1^st^ March 2020 were due to confirmed COVID-19.

**Conclusions:** In this nationwide study, we show that 40% of 30-day deaths are due to non-cardiac causes. Non-cardiac deaths have increased even more from the start of the COVID-19 pandemic, with one in ten deaths from March 2020 being COVID-19 related. These findings raise a question of whether public reporting of PCI outcomes should be cause-specific.

## Introduction

Percutaneous coronary intervention (PCI) is the most common modality for coronary revascularization worldwide. (1,2) Advances in PCI techniques, stent platforms and operator experience have all led to a reduction in mortality following PCI in recent years. (3) (4) (5) Mortality post-PCI may be attributed to both cardiovascular (CV) causes and non-CV causes, although PCI mortality risk prediction models are focused on overall mortality. (6) (7) (8) This is particularly relevant in the context of public reporting of operator PCI outcomes, which is often perceived as a surrogate of operator skill and quality of health care provided, (9,10) even though 30-day mortality post PCI may not be directly related to the procedure. There are inconsistent findings amongst current data on the causes of death following PCI, with some studies suggesting that the majority of short-term mortality after PCI is CV in origin, while others showing that CV mortality represents a minority of all deaths. (7,11-13) However, the evidence to date is based on single or multicenter registry analyses, older procedural cohorts (e.g. pre 2010) and analyses from highly selected patient-groups (e.g. randomized controlled trials (RCTs)) that may not be generalizable to a contemporary PCI population. For example, a study of 115,191 patients undergoing PCI in the Veterans Affairs (VA) healthcare system showed that only a minority of deaths within 30 days were cardiac in origin (28%). (12) However, they analyzed a selected cohort comprising elderly males (median age 71 years, 99% males) that is not representative of national PCI practice across different healthcare systems. In contrast, an analysis of 21 RCT’s, CV mortality was at least 5-fold higher than non-CV mortality (0.5% vs. 0.1%) at 30 days. (7)

Data around changes in the cause of death from an all-comer national perspective following PCI are limited, particularly around the COVID-19 pandemic, that has infected more than 10 million patients worldwide, with the United Kingdom (UK) ranking second globally in terms of mortality.(14) Recent studies have demonstrated higher COVID-19 related mortality in patients with CV disease.(15-17) The underlying mechanisms behind increased COVID-19 mortality in patients with CV disease remain unclear, with possible explanations including factors such as advanced age, lower angiotensin converting enzyme (ACE)-2 levels and impaired immunity.(18) Little is known about the characteristics, as well as rates and causes of death, of patients undergoing PCI in the current COVID era, and how these compare to those before the pandemic. The present study was designed to examine national-level trends and causes of death, up to 30-days post-procedure, in a contemporary PCI cohort in England before and during the COVID-19 pandemic.

## Methods

### Data Source, Study Design and Population

All adults (aged ≥18 years) undergoing PCI between 1^st^ January 2017 and 10^th^ May 2020 in England were retrospectively analyzed from the British Cardiovascular Intervention Society (BCIS) registry, stratified by outcome into 4 groups according to outcome from the date of PCI: no death, in-hospital death, up to 30-day death post-discharge (excluding in-hospital death), and 30-day total death. There were no specified inclusion or exclusion criteria except missing data on death (299 cases), deaths occurring more than 30 days after PCI (n=12,220), multiple PCI procedures for the same patient (n=13,693), in which case only the final procedure was included with all other previous procedures excluded (flow diagram: **Supplementary Figure 1**). The BCIS registry comprises clinical, procedural and in-hospital outcome data for all procedures undertaken in the United Kingdom. (19,20) Mortality beyond the in-hospital phase was collected via record linkage with the Office for National Statistics (ONS) Civil Registrations of Death dataset (up to date as of 9^th^ June 2020). (21) The process of death certification and registration is a legal requirement in the United Kingdom where a doctor who has seen the deceased within the last 14 days of life must complete a Medical Cause of Death Certificate unless a post-mortem examination is planned. International Classification of Diseases, tenth revision (ICD-10) codes were used to extract data on the most prevalent primary causes of deaths (first line on the death certificate), including COVID-19, from the ONS Civil Registrations of Death dataset. A full list of the diagnosis codes used in the study is provided in **Supplementary Table 1**. Cardiac deaths included any death due to the following: ACS, heart failure, cardiogenic shock, cardiac arrest, pre-existing IHD and cardiac procedural complications.

### Outcomes

The main outcome was post-PCI death at specific time points: in-hospital, up to 30 days post-discharge, and 30-day total, stratified in to cardiac, non-cardiac and COVID-19 related.

### Statistical Analysis

For exploratory analysis, we examined patient and procedural characteristics of patients undergoing PCI according to their final outcome: no death, in-hospital death, up to 30-days post-discharge death and 30-day total death. Further comparisons were performed according to individual year, as well as pre- and post-COVID-19 pandemic in 2020 (procedures performed between 1^st^ January-29^th^ February, and 1^st^ March-10^th^ May, respectively), and clinical syndrome (stable angina vs. ACS). Age was normally distributed and, therefore summarized using mean and standard deviation (SD) and compared using the t-test. Categorical variables were summarized as percentages and analyzed using the chi squared (X^2^) test or Fisher’s exact test, where appropriate, and using the Kruskal-Wallis test for ordinal variables. Statistical analyses were performed using Stata 16 MP (College Station, TX). Multivariable logistic regression models were performed to examine predictors of cardiac, non-cardiac and COVID-related deaths, adjusting for the patient characteristics and procedural characteristics (only for cardiac and non-cardiac deaths) summarized in **Appendix A**, and are expressed as odds ratios (OR) with corresponding 95% confidence intervals (CI). Multiple imputation with chained equations was performed for variables with missing data prior to model fitting, with a total of 10 imputations. Model estimates were later combined using Rubin’s rules.(22) The frequency of missing data prior to imputation is provided in **Supplementary Table 2**.

### Ethical Approval

The UK Secretary of State for Health and Social Care has issued a time limited Notice under Regulation 3(4) of the NHS (Control of Patient Information Regulations) 2002 (COPI) to share confidential patient information. The study complies with the Declaration of Helsinki. This work was part of a work stream endorsed by the Scientific Advisory Group for Emergencies (SAGE), the body responsible for ensuring timely and coordinated scientific advice is made available to UK government decision makers. SAGE supports UK cross-government decisions in the Cabinet Office Briefing Room (COBR)) and by NHS England, which overseas commissioning decisions in the NHS, and NHS Improvement, which is responsible for overseeing quality of care in NHS hospitals.

## Results

A total of 273,141 patients underwent PCI between 1^st^ January 2017 and 10^th^ May 2020. Overall, procedural volumes were similar between 2017 to 2019 (∼83,000/year). The number of procedures per 100,000 population sharply declined between 1^st^ March 2020 to 10^th^ May 2020 compared with the same period in previous years (**Figure 1**) but was largely similar in January and February across all years.

**Figure 1.**
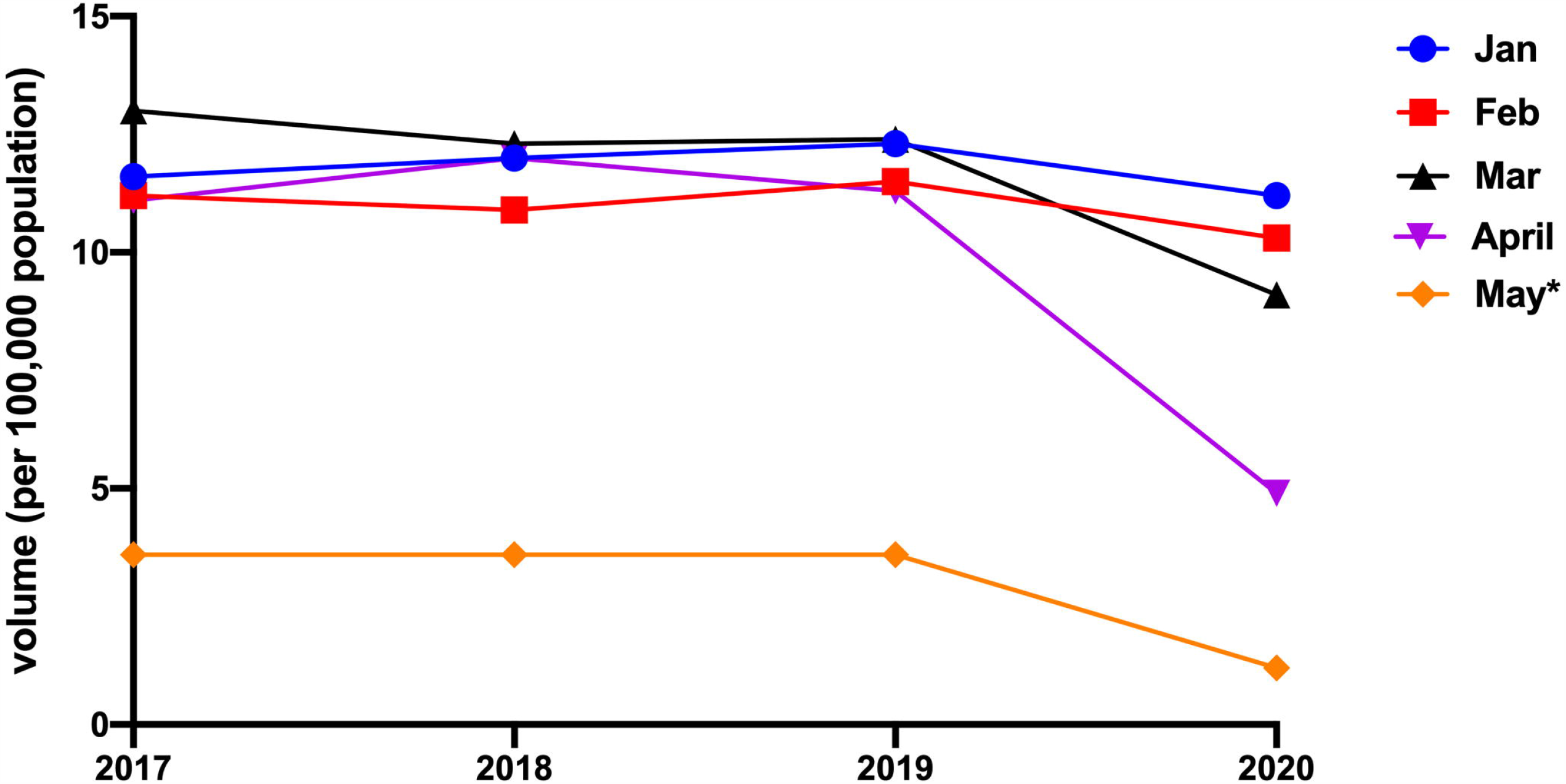
Trends of procedural volumes per 100,000 population over the study period. **Legend: \***Procedures until 10^th^ May 2020, p_trend_=non-significant for January-February, p_trend_ <0.001 for March to May

### i. 30-day death rates

The rate of 30-day death in the overall cohort was 2.8% (n=7,553), the majority of which occurred in hospital (1.9%, n=5,258). The rate of 30-day death declined between 2017 and February 2020 (2.9% to 2.5%, p<0.001), primarily driven by lower in-hospital death (2.1 vs. 1.5%), before rising again between 1^st^ March 2020 and 10^th^ May 2020 (3.2%) due to higher rates of post-discharge mortality up to 30 days. (**Figure 2A**) Overall, both in-hospital cardiac and non-cardiac death rates both declined over the study period (2017 to May 2020: cardiac: 1.21% vs. 1.00%; non-cardiac: 0.91% vs. 0.79%). Overall 59.7% (n=4,499) of 30-day deaths were due to cardiac causes. While 30-day cardiac and non-cardiac death rates both dropped between 2017 and February 2020, they were significantly increased in patients undergoing PCI between 1^st^ March and 10^th^ May 2020. (**Figure 2B**)

**Figure 2.**
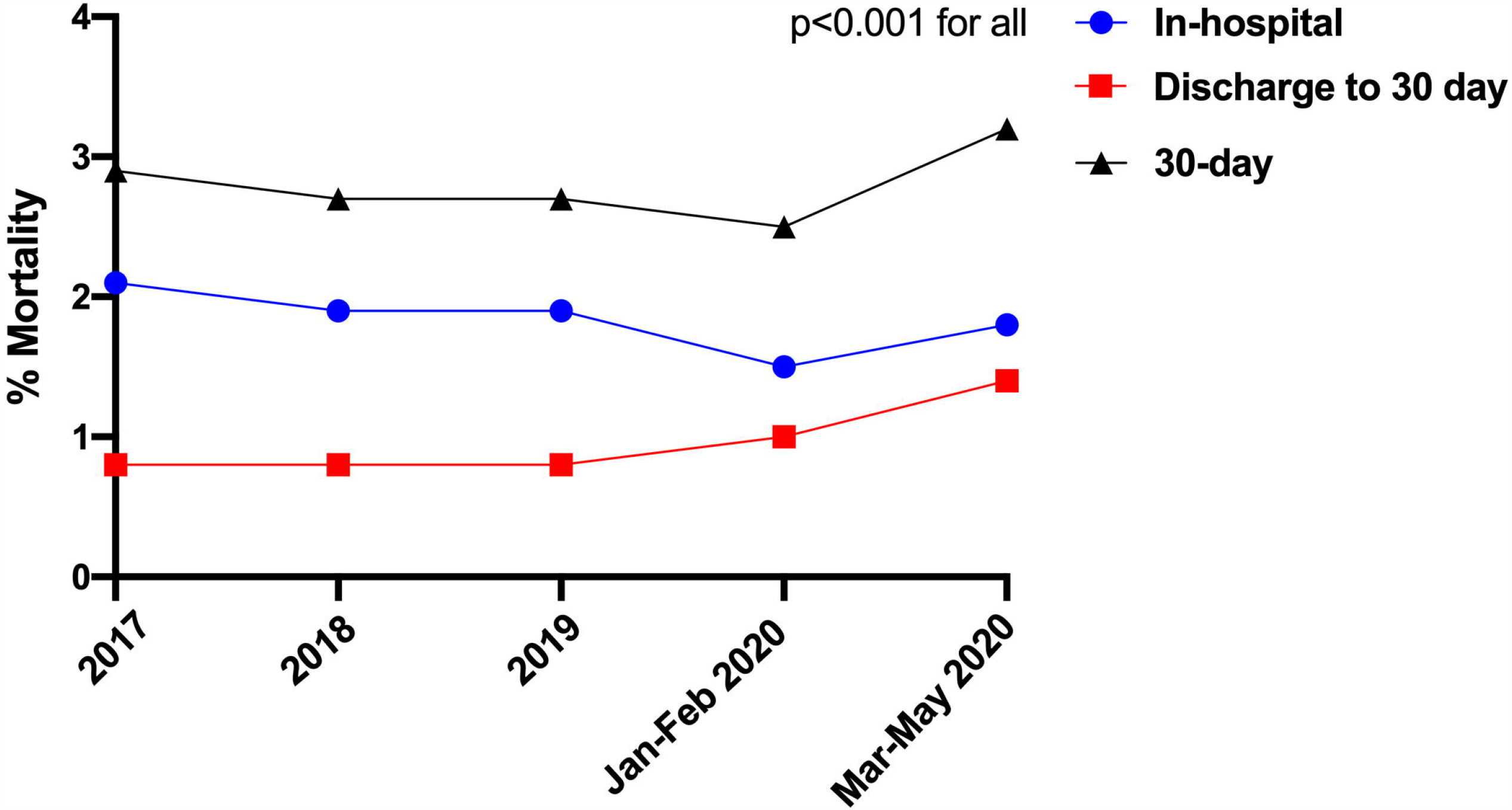
Trends of A) overall death and B) cardiac and non-cardiac death over the study period. **Legend: \***May procedures until 10^th^ May 2020

### ii. Patient and procedural characteristics

Overall, patients who died in-hospital or at 30-days were older (57 vs. 51 years), less likely to be males or from ethnic minorities, and more likely to have an ACS indication for their PCI. They had a greater prevalence of cardiovascular risk factors including previous coronary artery bypass grafts (CABG), diabetes, renal failure, moderate-poor left ventricular (LV) function, peripheral vascular disease (PVD) and previous cerebrovascular accidents (CVA). (**Table 1**) Furthermore, patients who died in-hospital and at 30-days were significantly more likely to have been in cardiogenic shock or suffered cardiac arrest or received mechanical ventilation prior to PCI. While this pattern of findings was consistent across the study years, patients undergoing PCI in 2020, particularly between March and May, were significantly younger (41-47 years vs. 51-57 years). (**Table 2**)

**Table 1.**
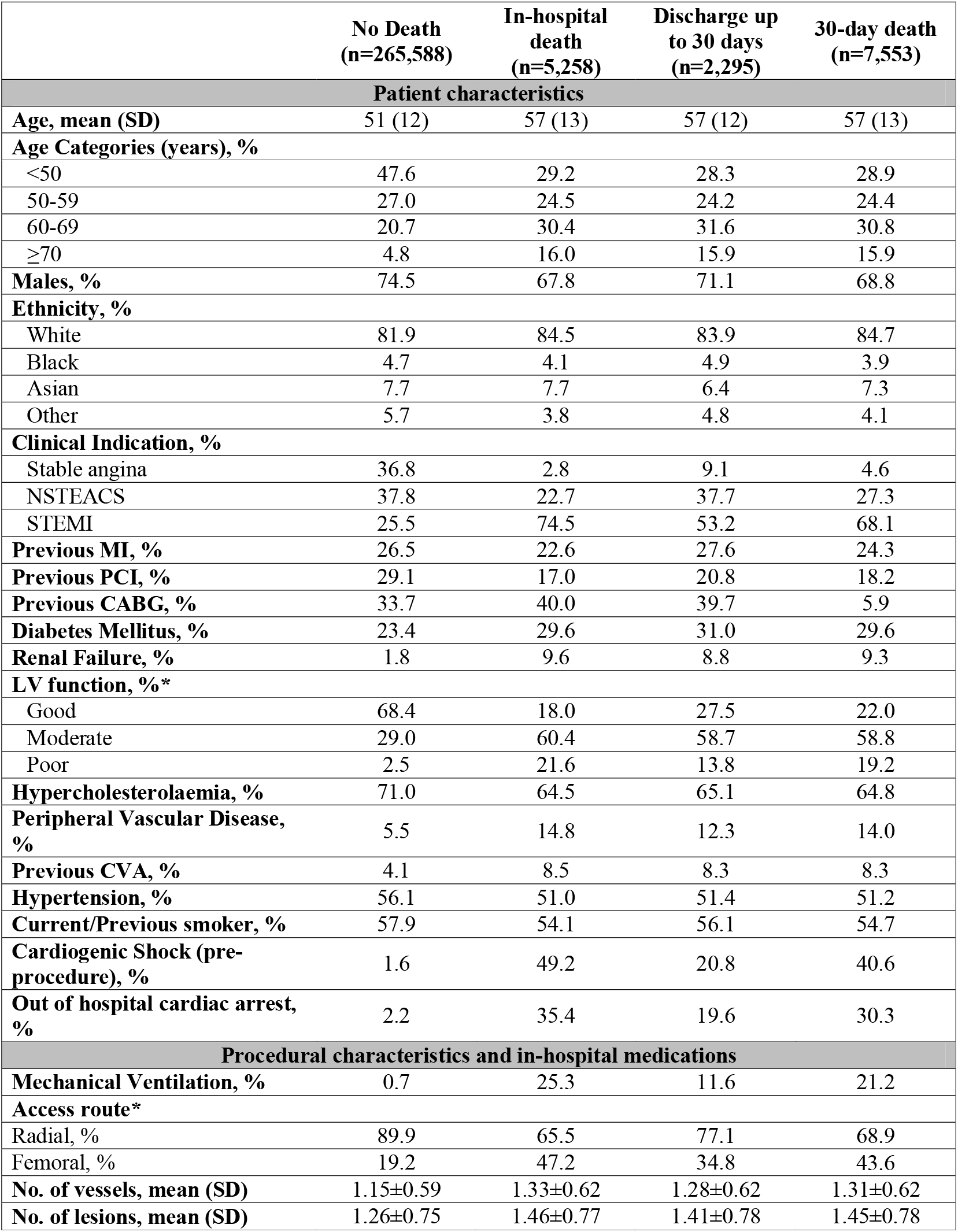
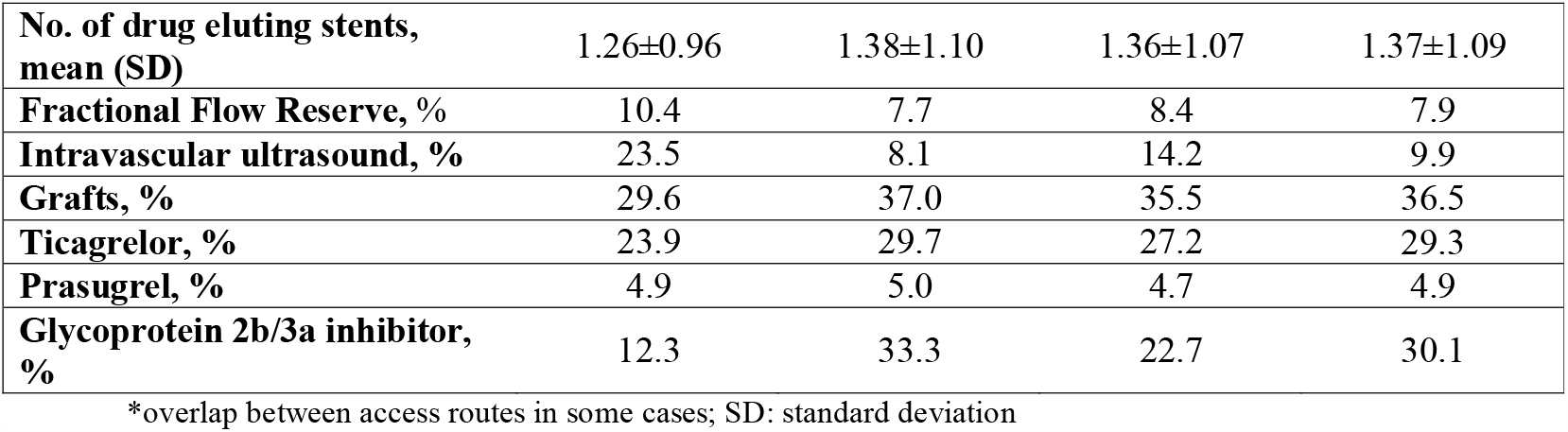
Patient and procedural characteristics of the total cohort.

**Table 2.** Patient and procedural characteristics of study groups according to year.

|  | 2017-2019<br>(n=251,304) |  |  |  | Jan-Feb 2020<br>(n=12,774) |  |  |  | Mar-May 2020<br>(n=9,063) |  |  |  |
| --- | --- | --- | --- | --- | --- | --- | --- | --- | --- | --- | --- | --- |
|  | No Death | In-hospital | Discharge to 30-days | 30-day total | No Death | In-hospital | Discharge to 30-days | 30-day total | No Death | In-hospital | Discharge to 30-days | 30-day total |
| **Number (% within group)** | 244353<br>(97.2) | 4915<br>(2.0) | 2036<br>(0.8) | 6951<br>(2.8) | 12458<br>(97.5) | 184<br>(1.5) | 132<br>(1.1) | 316<br>(2.6) | 8777<br>(96.8) | 159<br>(1.8) | 127<br>(1.4) | 286<br>(3.2) |
| **Patient characteristics** |  |  |  |  |  |  |  |  |  |  |  |  |
| **Age** | 51 (12) | 57 (13) | 57 (12) | 57 (13) | 41 (12) | 47 (12) | 47 (11) | 47 (12) | 40 (12) | 46 (11) | 46 (12) | 46 (11) |
| **Males, %** | 74.4 | 67.7 | 70.9 | 68.6 | 74.5 | 65.8 | 72.7 | 68.7 | 75.2 | 73.6 | 73.2 | 73.8 |
| **Ethnicity, %** |  |  |  |  |  |  |  |  |  |  |  |  |
| White | 81.8 | 84.8 | 84.7 | 84.8 | 82.2 | 85.3 | 78.8 | 84.2 | 82.8 | 83.6 | 82.7 | 83.6 |
| Black | 4.8 | 3.8 | 4.3 | 3.9 | 5.1 | 2.7 | 6.8 | 2.8 | 4.6 | 4.4 | 6.3 | 4.9 |
| Asian | 7.7 | 7.6 | 6.2 | 7.2 | 7.7 | 7.6 | 9.1 | 8.2 | 7.7 | 8.8 | 6.3 | 7.7 |
| Other | 5.8 | 3.8 | 4.8 | 4.1 | 4.9 | 4.3 | 5.3 | 4.7 | 5.0 | 3.1 | 4.7 | 3.8 |
| **Clinical Indication, %** |  |  |  |  |  |  |  |  |  |  |  |  |
| Stable angina | 37.1 | 2.6 | 9.3 | 4.5 | 36.1 | 4.3 | 6.8 | 5.1 | 28.7 | 3.1 | 9.4 | 5.6 |
| NSTEACS | 37.6 | 23.1 | 37.4 | 27.2 | 38.4 | 20.7 | 38.6 | 28.2 | 41.8 | 14.5 | 42.5 | 26.9 |
| STEMI | 25.3 | 74.4 | 53.3 | 68.2 | 25.4 | 75.0 | 54.5 | 66.8 | 29.5 | 82.4 | 48.0 | 67.5 |
| **Previous MI, %** | 26.6 | 23.2 | 27.5 | 24.4 | 26.3 | 17.9 | 29.5 | 23.1 | 24.7 | 17.0 | 28.3 | 22.0 |
| **Previous PCI, %** | 29.0 | 17.2 | 20.4 | 18.2 | 30.4 | 15.2 | 25.8 | 19.6 | 27.7 | 15.1 | 22.0 | 18.5 |
| **Previous CABG, %** | 6.7 | 5.4 | 7.1 | 5.9 | 33.9 | 45.7 | 43.2 | 6.3 | 33.2 | 39.0 | 38.6 | 4.5 |
| **Diabetes Mellitus, %** | 23.3 | 29.0 | 30.6 | 29.5 | 24.7 | 29.3 | 28.8 | 28.8 | 24.0 | 30.8 | 29.9 | 32.2 |
| **Renal Failure, %** | 1.8 | 9.5 | 8.7 | 9.3 | 2.2 | 9.3 | 7.4 | 7.6 | 2.0 | 8.1 | 13.3 | 11.2 |
| **LV function, %\*** |  |  |  |  |  |  |  |  |  |  |  |  |
| Good | 68.6 | 18.5 | 29.7 | 21.8 | 68.5 | 19.0 | 18.2 | 22.5 | 65.9 | 17.6 | 35.4 | 25.5 |
| Moderate | 28.9 | 59.9 | 56.1 | 58.8 | 28.7 | 58.2 | 69.7 | 59.2 | 31.5 | 61.6 | 55.1 | 58.7 |
| Poor | 2.5 | 21.6 | 14.1 | 19.4 | 2.7 | 22.8 | 12.1 | 18.4 | 2.7 | 20.8 | 9.4 | 15.7 |
| **Hypercholesterolaemia, %** | 71.0 | 64.9 | 64.1 | 64.7 | 70.9 | 62.0 | 72.0 | 64.9 | 69.3 | 62.9 | 72.4 | 67.8 |
| **Peripheral Vascular Disease, %** | 5.5 | 14.9 | 12.1 | 14.1 | 5.8 | 14.1 | 15.9 | 13.9 | 5.4 | 13.2 | 10.2 | 11.9 |
| **Previous CVA, %** | 4.4 | 8.6 | 8.7 | 8.6 | 1.5 | 4.9 | 4.5 | 3.8 | 1.5 | 4.4 | 3.9 | 4.2 |
| **Hypertension, %** | 56.1 | 51.1 | 50.8 | 51.0 | 57.0 | 51.6 | 56.1 | 52.2 | 55.6 | 49.7 | 59.8 | 54.2 |
| **Previous/current smoker, %** | 58.0 | 54.3 | 56.4 | 54.9 | 56.8 | 52.2 | 50.0 | 51.3 | 56.0 | 50.3 | 58.3 | 53.8 |
| **Cardiogenic Shock (pre-procedure), %** | 1.6 | 49.3 | 20.7 | 40.9 | 1.6 | 41.8 | 23.5 | 34.2 | 1.7 | 54.1 | 20.5 | 39.2 |
| **Out of hospital cardiac arrest, %** | 2.2 | 35.5 | 19.1 | 30.7 | 2.1 | 32.6 | 20.5 | 28.5 | 2.0 | 30.2 | 14.2 | 22.7 |
| **Procedural characteristics and in-hospital medications** |  |  |  |  |  |  |  |  |  |  |  |  |
| **Mechanical Ventilation, %** | 0.7 | 25.4 | 11.7 | 21.4 | 0.8 | 23.9 | 12.9 | 19.3 | 0.6 | 26.4 | 7.9 | 18.2 |
| **Access route\*** |  |  |  |  |  |  |  |  |  |  |  |  |
| Radial, % | 89.7 | 65.0 | 77.2 | 68.6 | 91.2 | 69.0 | 75.0 | 71.2 | 92.1 | 74.2 | 74.8 | 74.1 |
| Femoral, % | 19.3 | 48.2 | 35.4 | 44.5 | 19.6 | 37.5 | 36.4 | 37.3 | 17.9 | 31.4 | 27.6 | 30.1 |
| **No. of vessels, mean (SD)** | 1.14±0.5<br>9 | 1.33±0.6<br>2 | 1.27±0.6<br>2 | 1.31±0.6<br>2 | 1.14±0.5<br>9 | 1.28±0.5<br>7 | 1.25±0.5<br>9 | 1.27±0.5<br>8 | 1.19±0.6<br>1 | 1.31±0.6<br>2 | 1.35±0.7<br>0 | 1.33±0.6<br>5 |
| **No. of lesions, mean (SD)** | 1.26±0.7<br>5 | 1.47±0.7<br>7 | 1.42±0.7<br>9 | 1.45±0.7<br>9 | 1.23±0.7<br>3 | 1.41±0.7<br>3 | 1.33±0.7<br>6 | 1.38±0.7<br>4 | 1.31±0.7<br>6 | 1.43±0.7<br>7 | 1.48±0.8<br>1 | 1.45±0.7<br>8 |
| **No. of drug eluting stents, mean (SD)** | 1.26±0.9<br>6 | 1.39±1.1<br>0 | 1.37±1.0<br>9 | 1.38±1.1<br>0 | 1.19±0.9<br>2 | 1.23±1.0<br>7 | 1.25±0.9<br>9 | 1.25±1.0<br>4 | 1.26±0.9<br>5 | 1.27±1.0<br>3 | 1.39±0.9<br>8 | 1.32±1.0<br>1 |
| **Fractional Flow Reserve, %** | 10.4 | 7.5 | 8.6 | 7.9 | 10.1 | 9.2 | 9.1 | 9.8 | 10.0 | 10.1 | 3.1 | 6.6 |
| **Intravascular ultrasound, %** | 23.4 | 7.9 | 14.3 | 9.8 | 26.9 | 10.3 | 15.2 | 12.3 | 22.9 | 11.9 | 11.0 | 11.5 |
| **Grafts, %** | 29.8 | 36.8 | 35.2 | 36.4 | 29.7 | 41.3 | 40.2 | 41.1 | 30.0 | 38.4 | 32.3 | 35.0 |
| **Ticagrelor, %** | 24.9 | 30.6 | 29.2 | 30.2 | 12.1 | 22.8 | 15.2 | 21.5 | 12.1 | 14.5 | 13.4 | 14.7 |
| **Prasugrel, %** | 5.1 | 5.2 | 4.6 | 5.0 | 2.0 | 3.8 | 3.0 | 2.2 | 2.1 | 1.9 | 3.1 | 5.9 |
| **Glycoprotein 2b/3a inhibitor, %** | 12.2 | 33.2 | 22.9 | 30.2 | 11.9 | 32.1 | 23.5 | 28.5 | 13.6 | 37.1 | 19.7 | 29.4 |

\*overlap between access routes in some cases; SD: standard deviation

In terms of procedural characteristics, patients who died in-hospital or at 30-days were more likely to undergo a procedure via femoral access (34.8-47.2% vs. 19.2%), for grafts (35.5-37% vs. 29.6%) and multi-vessel disease (mean: 1.28-1.33 vs. 1.15) as well as multiple lesions (mean:: 1.41-1.46 vs. 1.25), using a greater number of drug eluting stents (DES; mean: 1.36-1.38 vs. 1.26) compared with those who did not die. (**Table 1**) Furthermore, they were less likely to undergo intravascular ultrasound (IVUS; 8.1-14.2% vs. 23.5%) and fractional flow reserve assessment (FFR’ 7.7-8.4% vs. 10.4%), and more likely to receive glycoprotein 2b/3a inhibitors (22.7-33.3% vs. 12.3%). Patients who died within 30 days were also more likely to receive ticagrelor (27.2-29.7% vs. 23.9%), but no difference in the utilization of prasugrel was observed between groups. These findings persisted throughout the study period (**Table 2**).

### iii. 30-day causes of death

One in ten deaths (10.4%) at 30 days were due do confirmed COVID in those who underwent PCI between 1^st^ March and 10^th^ May 2020. (**Table 3A**) Overall, the majority of deaths within 30 days were cardiovascular in origin (59.6%, n=4,499), and this persisted between 2017 and 2020. (**Figure 3**) Cardiac deaths were mostly due to ACS (33.2%) and cardiogenic shock (10.2%), more so for in-hospital deaths than post-discharges, followed by heart failure (7.5%) which was higher in post-discharge than in-hospital deaths. (30-day death illustrated in **Figure 4**) While this pattern was consistent over the study period, the rate of in-hospital death due to cardiogenic shock significantly increased between 1^st^ March and 10^th^ May 2020 (17.6% vs. 9.4-12.8%). (**Table 3B**) The most common causes of non-cardiac death were hypoxic brain injury for in-hospital death (10.9%) and non-COVID infections for post-discharge death (4.2%). (**Table 3A**) Deaths due to infections (non-COVID) increased in 2020, both in-hospital and post-discharge, whereas deaths due to hypoxic brain injury declined over the same period, particularly post-discharge. (**Table 3B**)

**Table 3A.**
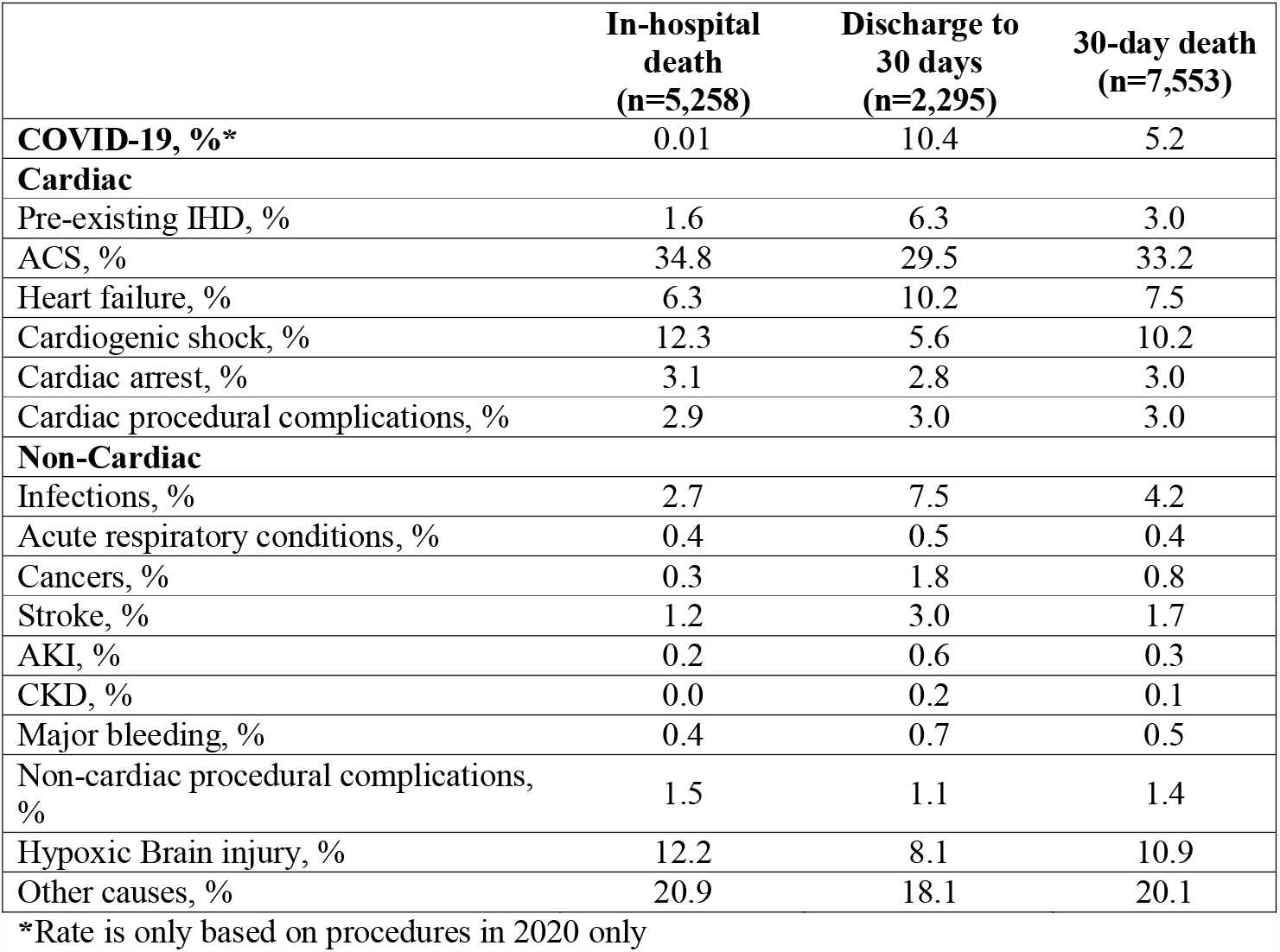
In-hospital and 30-day primary cause of death for the total cohort.

**Table 3B.** In-hospital and 30-day primary cause of death according to year.

|  | 2017-2019<br>(n=6,951) |  |  | Jan-Feb 2020<br>(n=316) |  |  | Mar-May 2020<br>(n=286) |  |  |
| --- | --- | --- | --- | --- | --- | --- | --- | --- | --- |
|  | In-hospital<br>(n=4915) | Discharge to 30-days<br>(n=2036) | 30-day<br>total | In-hospital<br>(n=184) | Discharge to 30-days<br>(n=132) | 30-day<br>total | In-hospital<br>(n=159) | Discharge to 30-days<br>(n=127) | 30-day<br>total |
| **COVID-19, %** | - | - | - | 0.0 | 1.5 | 0.01 | 2.0 | 19.7 | 10.2 |
| **Cardiac** |  |  |  |  |  |  |  |  |  |
| Pre-existing IHD, % | 1.5 | 6.5 | 3.0 | 0.5 | 5.3 | 2.5 | 0.0 | 1.6 | 0.7 |
| ACS, % | 34.2 | 29.3 | 32.7 | 41.3 | 32.6 | 37.7 | 32.7 | 35.4 | 33.9 |
| Heart failure, % | 6.3 | 10.4 | 7.5 | 5.4 | 8.3 | 6.7 | 6.9 | 7.9 | 7.3 |
| Cardiogenic shock, % | 12.8 | 5.9 | 10.7 | 9.4 | 12.1 | 10.6 | 17.6 | 4.7 | 11.6 |
| Cardiac arrest, % | 2.9 | 2.6 | 2.8 | 2.4 | 6.1 | 4.0 | 2.7 | 1.6 | 2.2 |
| Cardiac procedural complications, % | 3.1 | 3.4 | 3.2 | 0.6 | 0.0 | 0.3 | 0.7 | 0.0 | 0.4 |
| **Non-cardiac** |  |  |  |  |  |  |  |  |  |
| Infections, % | 2.5 | 6.9 | 3.8 | 4.9 | 6.8 | 5.7 | 5.0 | 9.4 | 7.0 |
| Acute respiratory conditions, % | 0.4 | 0.6 | 0.5 | - | - | - | 0.0 | 1.6 | 0.7 |
| Cancers, % | 0.3 | 1.4 | 0.6 | 0.0 | 1.5 | 0.6 | 1.3 | 0.8 | 1.0 |
| Stroke, % | 1.2 | 2.9 | 1.7 | 0.5 | 1.5 | 0.9 | 1.3 | 1.6 | 1.4 |
| AKI, % | 0.1 | 0.9 | 0.4 | - | - | - | - | - | - |
| Major bleeding, % | 0.3 | 0.2 | 0.2 | 0.0 | 0.8 | 0.3 | - | - | - |
| Non-cardiac procedural complications, % | 1.6 | 1.2 | 1.5 | 0 | 0 | 0 | 0 | 0 | 0 |
| Hypoxic Brain injury, % | 12.3 | 8.5 | 11.2 | 10.6 | 8.3 | 9.6 | 10.8 | 1.6 | 6.6 |
| Other causes, % | 20.7 | 18.5 | 20.1 | 26.0 | 15.2 | 21.5 | 21.4 | 14.2 | 18.2 |

**Figure 3.**
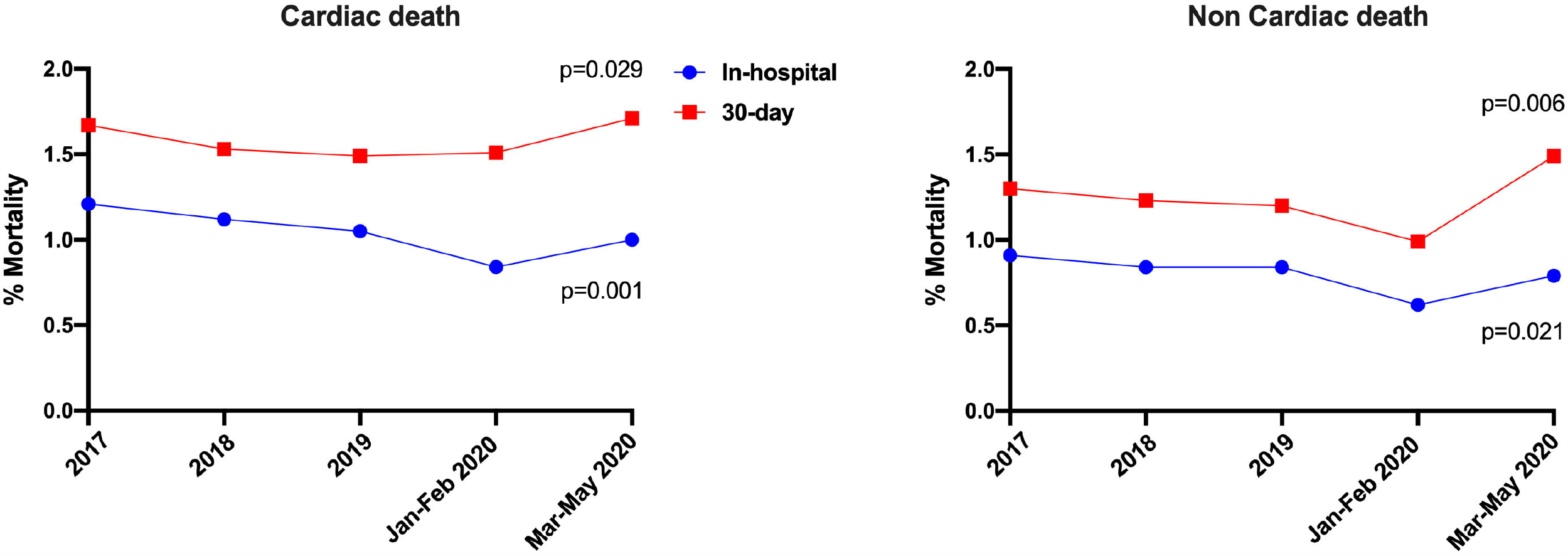
Figure 3. Trends of 30-day causes of death over the study period.

**Figure 4.**
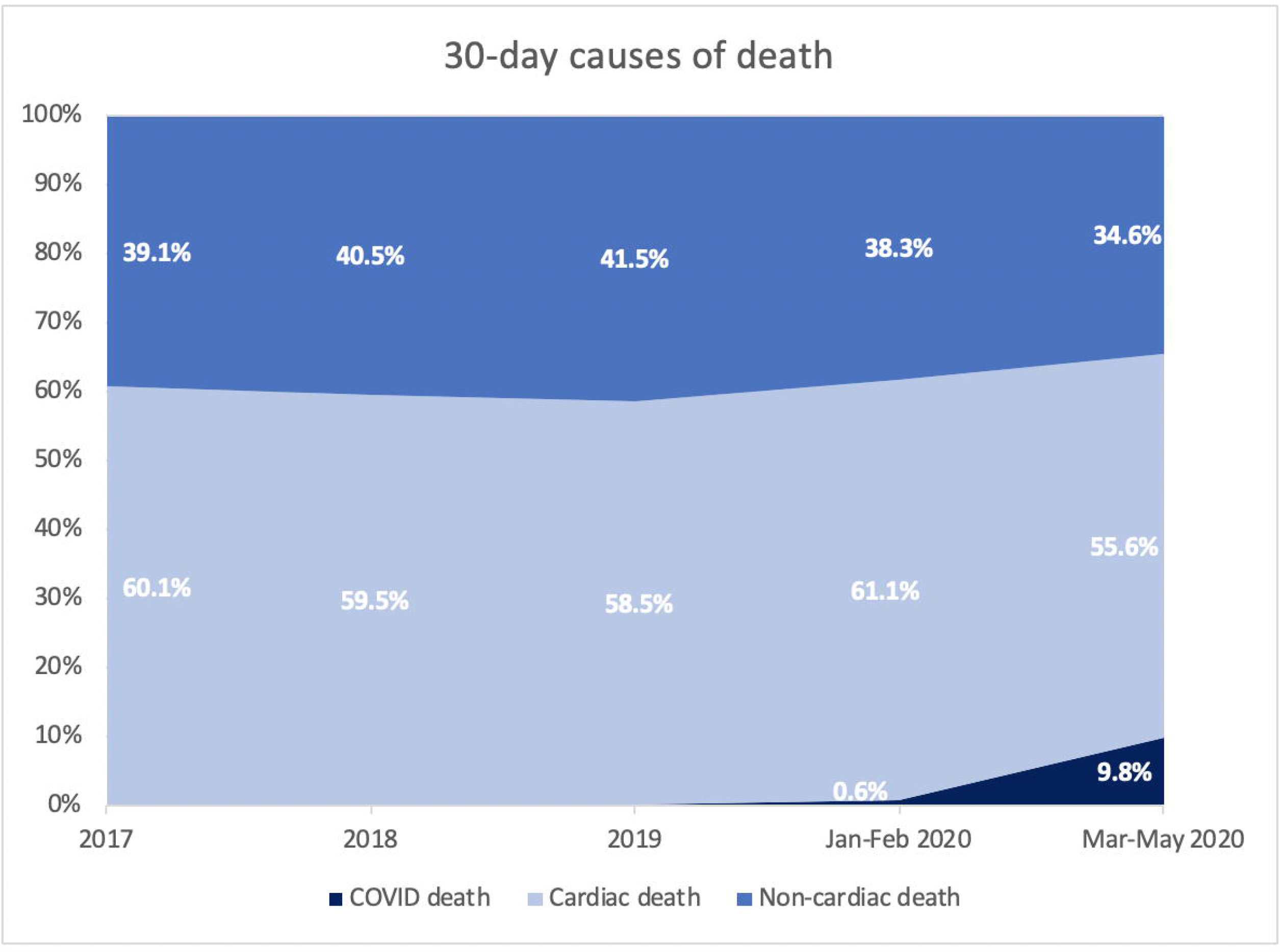
Figure 4. Causes of 30-day mortality in A)2017-2019, B) January-February 2020 and C) March-May 2020*. **Legend:** *Correct as of 9th June 2020

When stratified by PCI indication, the majority of 30-day deaths occurred in patients who underwent PCI for ACS (n=7,205; 95.4% of all deaths). Within this group, the most common cause of 30-day death was cardiac in origin (60.2%), with a third of deaths primarily due to ACS (33.9%). Hypoxic brain injury and cardiogenic shock were the most prevalent other causes (11.3% and 10.6%, respectively), all of which were more likely to contribute to in-hospital than post-discharge death. (**Supplementary Table 3**) In patients who underwent PCI for stable angina, the most common causes of 30-day death were non-cardiac (53.6%). The most common cardiac causes of 30-day death in the stable angina group were ACS (17.2%), heart failure (14.1%) and pre-existing heart disease (8.6%).

### iv. Predictors of death

Several factors correlated with increased odds of both 30-day cardiac and non-cardiac, including advanced age (>50 years), ACS compared with stable angina (STEMI>NSTEACS), adverse events prior to PCI including cardiogenic shock, out-of-hospital cardiac arrest and need for pre-PCI mechanical ventilation, renal failure, moderate-poor left ventricular function, PVD, diabetes mellitus and history of MI or CVA. (**Table 4**) Amongst the procedural characteristics, the odds of 30-day cardiac death were increased with femoral access (OR 1.62 95% CI 1.47, 1.77), graft PCI (OR 1.24 95% CI 1.16, 1.32) and greater number of lesions (per additional lesion: OR 1.15 95% CI 1.06, 1.23) or vessels (per additional vessel: OR 1.34 95% CI 1.23, 1.46) (p<0.001 for all). In contrast, the odds of 30-day cardiac and non-cardiac death were lower amongst males (OR 0.70 95% CI 0.66, 0.75 and 0.77 95% CI 0.72, 0.83, respectively, p<0.001 for both) and non-white ethnic backgrounds. (**Table 4**) Furthermore, the odds of 30-day cardiac death were reduced with radial access (OR 0.78 95% CI 0.70, 0.86) and the use of IVUS (OR 0.75 95% CI 0.67, 0.83), FFR (OR 0.81 95% CI 0.72, 0.91) and DES (OR 0.78 95% CI 0.68, 0.89) (p<0.001 for all). The odds of COVID-related 30-day death were significantly increased in patients aged 60-69 years (OR 5.61 95% CI 2.07, 15.16, p=0.001) and those with renal failure (OR 7.31 95% CI 2.66, 20.06, p<0.001). Although there was a trend towards increased COVID-related death amongst Black and Asian ethnicities (OR 2.95 95% CI 0.98, 8.84 and 1.70 95% CI 0.55, 5.26, respectively) and males (OR 1.59 95% CI 0.60, 4.23), this was not statistically significant, possibly due to the relatively small number of COVID-19 deaths.

**Table 4.** Predictors of in-hospital and 30-day cardiac, non-cardiac and COVID-related deaths.

| Predictor/<br>Outcome | 30-day cardiac |  | 30-day non-cardiac |  | 30-day COVID |  |
| --- | --- | --- | --- | --- | --- | --- |
|  | OR (95% CI) | P-value | OR (95% CI) | P-value | OR (95% CI) | P-value |
| **Patient characteristics** |  |  |  |  |  |  |
| **Age category (years)** |  |  |  |  |  |  |
| <50 (reference) |  |  |  |  |  |  |
| 50-59 | 1.98 [1.79, 2.19] | <0.001 | 1.71 [1.54, 1.89] | <0.001 | 1.73 [0.54, 5.52] | 0.352 |
| 60-69 | 3.44 [3.12, 3.79] | <0.001 | 2.68 [2.42, 2.98] | <0.001 | 5.61 [2.07, 15.16] | 0.001 |
| ≥70 | 7.36 [6.55, 8.27] | <0.001 | 4.47 [3.90, 5.12] | <0.001 | 3.39 [0.64, 17.92] | 0.150 |
| **Male** | 0.70 [0.66, 0.75] | <0.001 | 0.77 [0.72, 0.83] | <0.001 | 1.59 [0.60, 4.23] | 0.353 |
| **Race** |  |  |  |  |  |  |
| White (reference) |  |  |  |  |  |  |
| Black | 0.75 [0.63, 0.88] | <0.001 | 0.87 [0.74, 1.02] | 0.094 | 2.95 [0.98, 8.84] | 0.054 |
| Asian | 0.86 [0.76, 0.97] | 0.016 | 0.86 [0.75, 0.98] | 0.025 | 1.70 [0.55, 5.26] | 0.355 |
| Other | 0.82 [0.71, 0.96] | 0.015 | 0.88 [0.75, 1.03] | 0.099 | 0.74 [0.10, 5.64] | 0.775 |
| **Clinical Syndrome** |  |  |  |  |  |  |
| Stable angina (reference) |  |  |  |  |  |  |
| NSTEACS | 3.78 [3.30, 4.34] | <0.001 | 3.17 [2.79, 3.61] | <0.001 | 2.98 [0.97, 9.15] | 0.056 |
| STEMI | 7.45 [6.46, 8.58] | <0.001 | 5.17 [4.51, 5.92] | <0.001 | 2.32 [0.65, 8.24] | 0.193 |
| **Previous MI** | 1.19 [1.08, 1.30] | <0.001 | 1.12 [1.01, 1.24] | 0.026 | 0.82 [0.30, 2.22] | 0.691 |
| **Previous PCI** | 0.65 [0.58, 0.71] | <0.001 | 0.73 [0.66, 0.81] | <0.001 | 1.80 [0.70, 4.63] | 0.222 |
| **Previous CABG** | 0.80 [0.70, 0.92] | 0.001 | 0.86 [0.75, 0.99] | 0.033 | 1.28 [0.36, 4.58] | 0.706 |
| **Diabetes Mellitus** | 1.49 [1.39, 1.60] | <0.001 | 1.39 [1.29, 1.50] | <0.001 | 1.15 [0.51, 2.61] | 0.739 |
| **Renal Failure** | 3.77 [3.33, 4.25] | <0.001 | 3.99 [3.52, 4.52] | <0.001 | 7.31 [2.66, 20.06] | <0.001 |
| **LV Function** |  |  |  |  |  |  |
| Good (reference) |  |  |  |  |  |  |
| Moderate | 2.65 [2.46, 2.86] | <0.001 | 2.37 [2.19, 2.55] | <0.001 | 1.41 [0.64, 3.10] | 0.397 |
| Poor | 6.41 [5.77, 7.13] | <0.001 | 5.12 [4.57, 5.74] | <0.001 | 1.06 [0.13, 8.70] | 0.957 |
| **Hypercholesterolaemia** | 0.95 [0.86, 1.04] | 0.230 | 0.96 [0.87, 1.05] | 0.349 | 1.12 [0.32, 3.88] | 0.862 |
| **PVD** | 1.54 [1.38, 1.72] | <0.001 | 1.50 [1.34, 1.68] | <0.001 | 1.64 [0.49, 5.51] | 0.420 |
| **Previous CVA** | 1.37 [1.20, 1.55] | <0.001 | 1.47 [1.28, 1.68] | <0.001 | 3.50 [0.64, 19.18] | 0.150 |
| **Hypertension** | 1.16 [1.06, 1.26] | 0.001 | 1.08 [0.98, 1.18] | 0.116 | 1.73 [0.56, 5.40] | 0.344 |
| **Cardiogenic Shock (pre-** | 8.39 [7.75, 9.07] | <0.001 | 5.23 [4.79, 5.71] | <0.001 | 2.74 [0.51, 14.62] | 0.238 |
| operative) |  |  |  |  |  |  |
| **Out-of-hospital arrest** | 1.80 [1.60, 2.02] | <0.001 | 3.06 [2.72, 3.43] | <0.001 | 1.69 [0.25, 11.52] | 0.590 |
| **Ventilation in-hospital** | 2.60 [2.25, 3.00] | <0.001 | 3.45 [3.01, 3.95] | <0.001 | 3.45 [0.25, 47.58] | 0.354 |
| **Medications and Procedural characteristics** |  |  |  |  |  |  |
| **P2Y12 inhibitor** | 1.00 [0.97, 1.03] | 0.918 | 0.98 [0.95, 1.01] | 0.250 | 1.26 [0.82,1.94] | 0.283 |
| **Glycoprotein 2b/3a inhibitor** | 0.92 [0.86, 1.00] | 0.038 | 0.91 [0.84, 0.98] | 0.017 |  |  |
| **Radial access** | 0.78 [0.70, 0.86] | <0.001 |  |  |  |  |
| **Femoral access** | 1.62 [1.47, 1.77] | <0.001 |  |  |  |  |
| **No. of vessels** | 1.34 [1.23, 1.46] | <0.001 |  |  |  |  |
| **No. of lesions** | 1.15 [1.06, 1.23] | <0.001 |  |  |  |  |
| **FFR** | 0.81 [0.72, 0.91] | <0.001 |  |  |  |  |
| **IVUS** | 0.75 [0.67, 0.83] | <0.001 |  |  |  |  |
| **Drug eluting stents** | 0.78 [0.68, 0.89] | <0.001 |  |  |  |  |
| **Grafts** | 1.24 [1.16, 1.32] | <0.001 |  |  |  |  |

## Discussion

The present study highlights several important findings from a contemporary nationwide cohort of patients treated by PCI in England. First, we find that only two of every three deaths at 30-days following PCI were from cardiac causes, and that this has persisted throughout the study period, even during the COVID-19 pandemic. Patients treated for an ACS accounted for a third of 30-day deaths throughout the study, followed by hypoxic brain injury and cardiogenic shock as the most prevalent causes. Second, we observe that in-hospital and 30-day deaths after PCI, from both cardiac and non-cardiac causes, have declined in the past three years, although these have risen again after the start of the COVID-19 pandemic (1^st^ March through 10^th^ May 2020), primarily due to non-cardiac deaths. One in ten deaths at 30 days were due to COVID-19 in patients undergoing PCI between March and May 2020.

Some previous studies have suggested that short-term mortality (≤30 days) after PCI is predominantly due to cardiac causes, while others have shown otherwise. (6,7,13) However, there are limited data on the causes of death after PCI in the current era, and how these have changed from a national perspective. Previous studies provided conflicting information, likely due to variations in the cohorts they had examined, which included individual centers (11) or healthcare systems (12), or only inpatient procedures, with no analysis of post-discharge causes of death.(5) Pooled data from 21 RCTs demonstrated a 7-fold higher rate of CV than non-CV deaths in 32,882 patients undergoing PCI (relative ratio: 6.99, 95% CI 3.16–15.42). (7) Despite the added value of this study, patients enrolled in RCTs are highly selected cohorts that are often healthier than the background population encountered in daily practice. Furthermore, the authors mention that death of unknown cause in RCTs was adjudicated as CV in origin as per protocol, which may have influenced the external validity of their findings. In contrast, a study by Bricker et al. reports higher rates (∼60%) of non-cardiovascular/undifferentiated deaths at 30-days in 115,191 patients undergoing PCI in the VA healthcare system in the US. However, their study examined a selected cohort of veterans, who were predominantly males (99%) and generally older than the average population. (12) Furthermore, deaths that occurred outside the VA were not captured, which may explain their apparently high number of non-cardiac deaths.

Although the majority of in-hospital and 30-day deaths in our cohort were cardiac in origin, 4 out of 10 deaths (∼40%) were non-cardiac, with hypoxic brain injury and infections being the most common non-cardiac causes, a finding that persisted over the study period. This finding raises questions regarding the utility of currently used PCI mortality risk scores, such as the New York State Risk (NYSR) score and National Cardiovascular Data Registry (NCDR) score. (6,8,23) While these scores have been validated for the prediction of overall in-hospital mortality, they do not discriminate between CV and non-CV deaths, with the latter representing a significant proportion of all in-hospital deaths. Furthermore, these risk scores were derived from relatively outdated cohorts (e.g. NCDR: 2004 to 2006; NYSR: 2009 to 2010), whose procedural characteristics are different to the current era. Similarly, an argument could be made that public reporting of operator outcomes should detail the broad nature of the cause of death, particularly in view of the cause of death distribution during the COVID-19 pandemic, where one in ten deaths at 30-days were due to COVID-19. Interestingly, most of the COVID-19 deaths in our cohort were amongst patients undergoing PCI for stable angina, although it does not determine whether the transmission of COVID-19 was nosocomial or post discharge in the community. Nevertheless, this raises a concern regarding the safety of performing non-emergent PCI until the pandemic has subsided.

There are limited data regarding trends in in-hospital and 30-day mortality in recent years. In a single-center study of 19,506 patients undergoing PCI in Mayo Clinic (Rochester, Minnesota), Spoon and colleagues reported a decline in in-hospital death rates between 1991 (2.7%) and 2006 (2.2%), as well as a reduction in long-term (5-year) cardiac death (33% temporal decline), with a parallel increased in long-term non-cardiac death (57%).(11) Their findings, however, were derived from a relatively old cohort that was less likely to receive DES and newer P2Y12 inhibitors, and one that was managed in a large tertiary facility and, thereby, does not reflect contemporary national-level practices or outcomes. Furthermore, their analysis does not provide information on the cause of in-hospital mortality (e.g. cardiac vs. non-cardiac), and only considered this aspect for long-term outcomes. In contrast, Alkhouli et al. reported an increase in in-hospital mortality after PCI between 2003 and 2016, both in ACS and stable angina patients, in a 20% stratified national sample of US hospitalizations.(5) While their analysis provides us with insights in to the trends of in-hospital mortality in inpatients undergoing PCI, it does not inform us on outcomes for outpatient procedures, which represent a large proportion of all PCI cases. Moreover, they did not report the cause of in-hospital mortality. Our findings suggest that both 30-day cardiac and non-cardiac death rates after PCI have declined over the past 3 years, mainly driven by a fall in in-hospital death, up until the start of the COVID-19 pandemic (1^st^ March 2020), after which there was an increase in both in-hospital cardiac and non-cardiac deaths. The latter could be explained by baseline differences between patients undergoing PCI before and after the pandemic. For example, patients undergoing PCI during the pandemic could have been more critically unwell, or higher-risk PCI cases, although we note that patient and procedural characteristics during both time periods were relatively similar. Another possible explanation is the delayed presentation of ACS cases during the pandemic, which have been widely reported in recent literature, as well as less optimal care for critically ill cases due to pressures on hospital systems as a result of the COVID-19 pandemic. (24,25) (26)

### Strengths and Limitations

The present study is the largest to report trends of 30-day causes of deaths after PCI in a contemporary nationwide cohort, including both inpatient and outpatient procedures, making it generalizable to the wider population of interest. Another notable strength is our ascertainment and determination of cause of death, which is based on linkage to the ONS database and details from the Medical Cause of Death Certificate, unlike some previous studies. However, there are several limitations to the present study. First, whilst the BCIS dataset captures cardiovascular risk factors and procedural characteristics, it does not capture measures of comorbidity, such as Charlson or Elixhauser scores, or frailty that are important determinants of mortality post PCI. (27,28) The observational nature of our study means that residual unmeasured confounders such as these and others may not be accounted for. Third, given that these data are the most contemporary available (census June 2020 for procedures in May 2020) there is insufficient follow up to study longer term causes of death, particularly given that it has been shown in patients derived from RCTs that non-cardiovascular causes of mortality become more important at longer term follow up.(7) Finally, COVID-19 cases were identified according to the corresponding ICD-10 code (U07.1 – confirmed COVID). However, the data does not inform us of whether this confirmation was based on virology/ serology or clinical diagnoses.

## Conclusions

In a nationwide cohort of PCI procedures, we demonstrate that a significant proportion of 30-day deaths is due to non-cardiac causes, a finding that has persisted over the past 3 years. Overall, in-hospital and 30-day cardiac and non-cardiac deaths have declined over 3 years, before increasing again during the COVID-19 pandemic. One in ten deaths in those undergoing PCI from 1^st^ March 2020 were due to COVID-19. The present findings highlight that overall 30-day mortality may not be a reliable measure of performance in the context of PCI, and drive the need for other objective measures, which may include cause-specific mortality.

## Data Availability

Data is not available for review as it is confidential and property of NHS Digital

## Acknowledgments

The authors acknowledge Chris Roebuck and Tom Denwood from NHS digital for providing and creating the secure environment for data hosting and for analytical support.

## Figures captions and legends

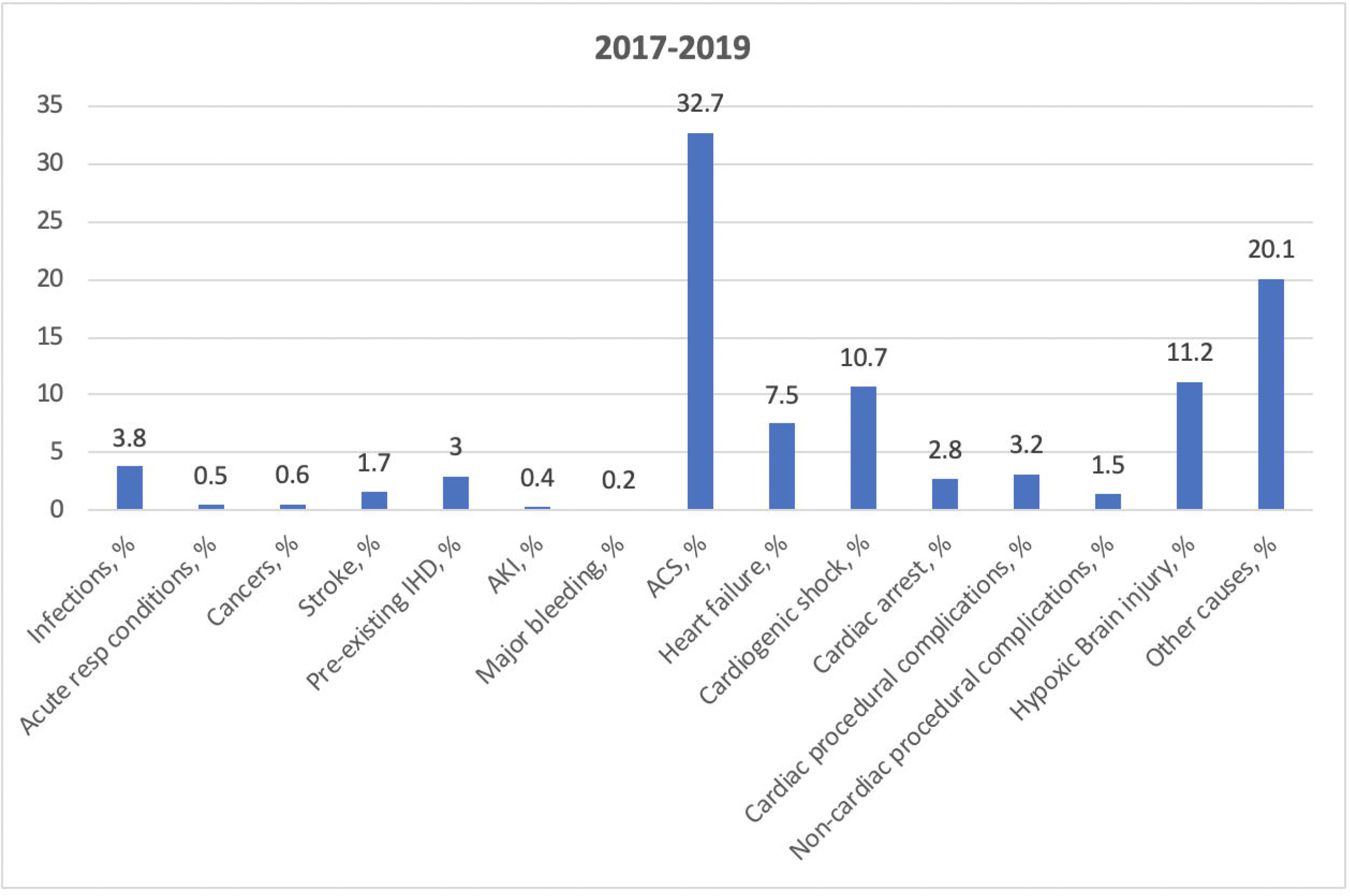

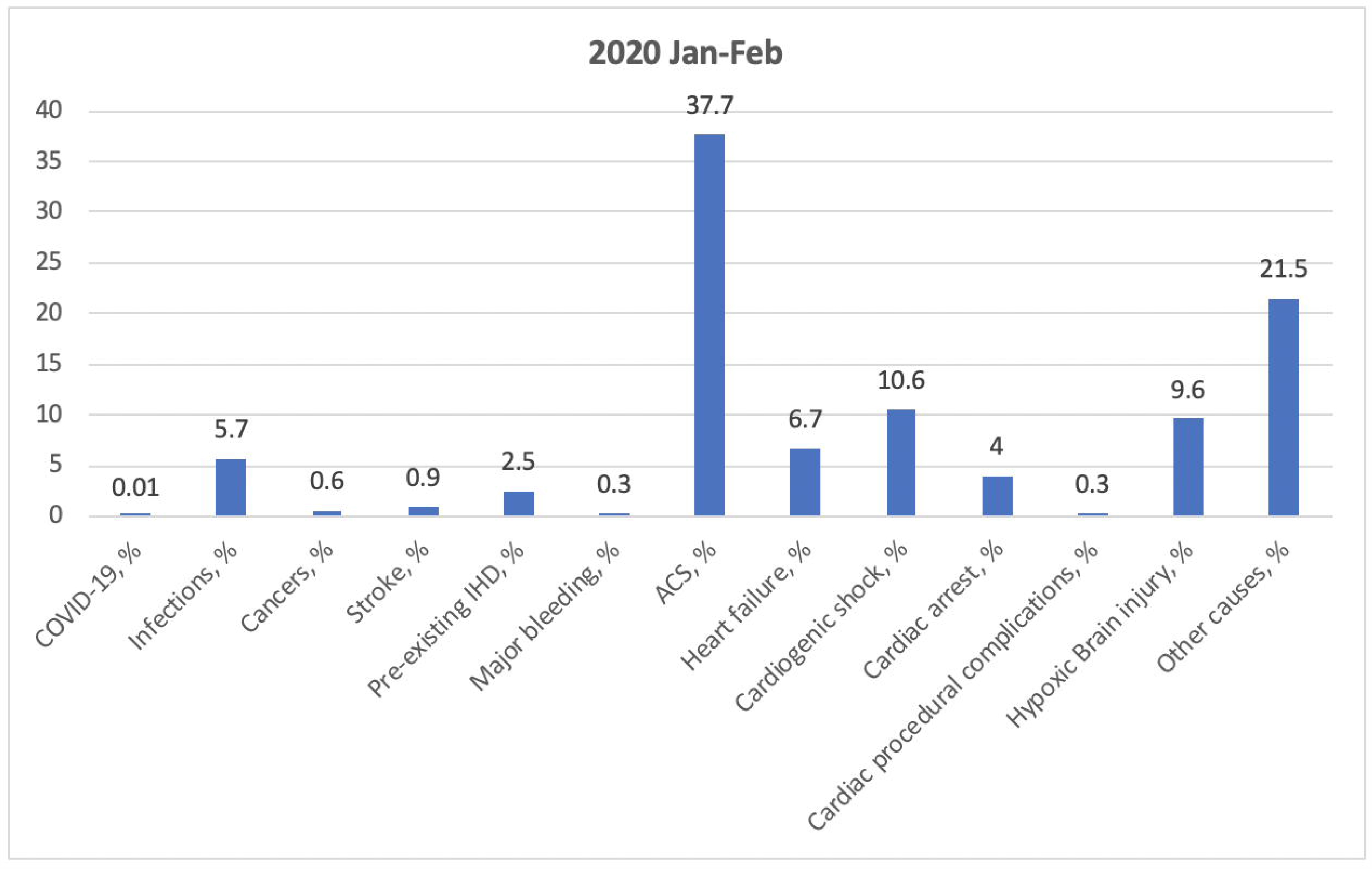

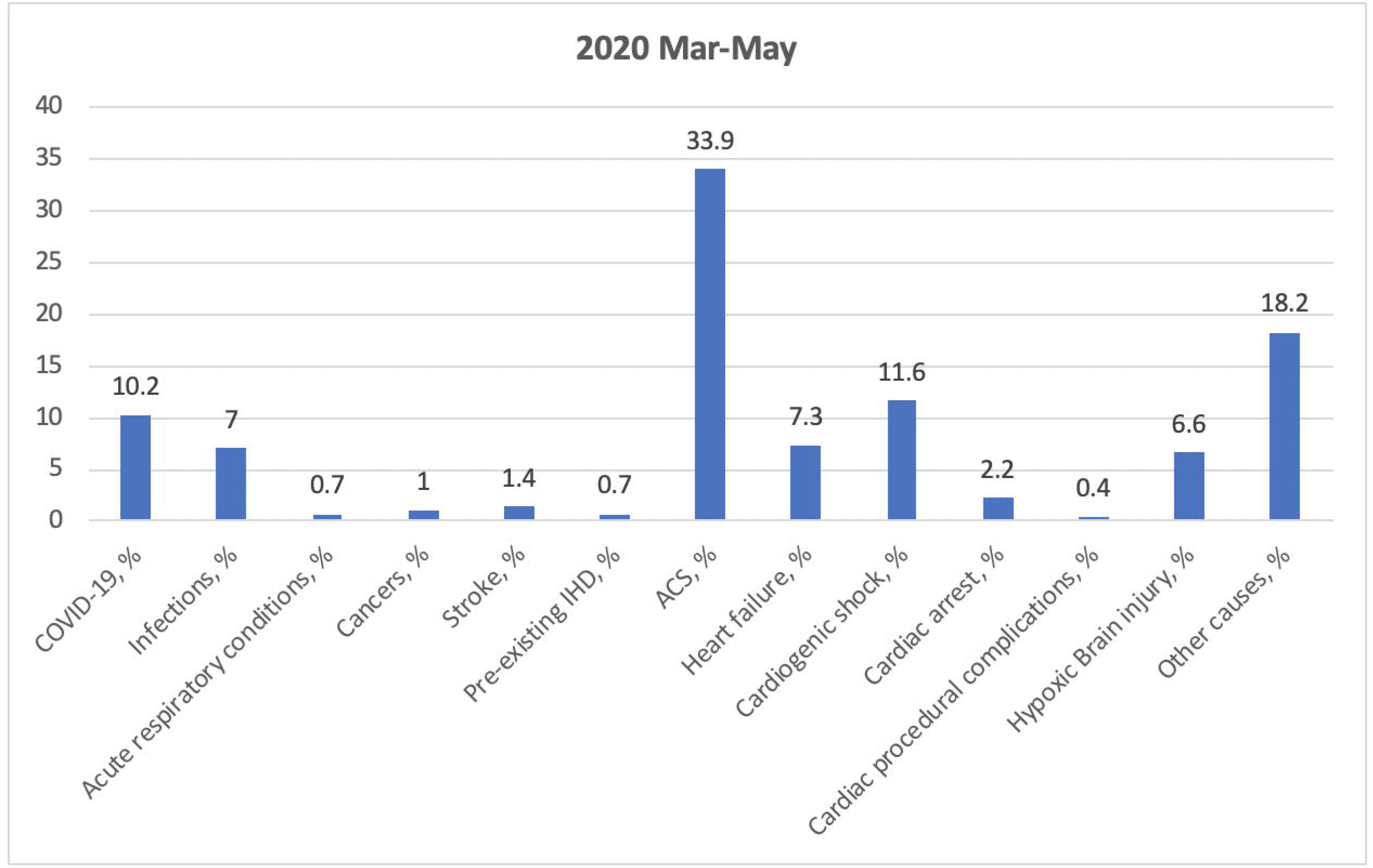

